# Temporal genome dynamics of ST39 *Klebsiella pneumoniae* in a neonatal unit in Blantyre, Malawi

**DOI:** 10.1101/2025.11.06.25339690

**Authors:** Allan M Zuza, Oliver Pearse, Daryl B Domman, Zoe A. Dyson, Kondwani Kawaza, Patrick Musicha, Nicholas A Feasey, Eva Heinz

## Abstract

**Background:** *Klebsiella pneumoniae* (*Kpn*) is an important cause of healthcare-associated infections (HAI). In low and middle-income countries, HAI due to *Kpn* disproportionally affects neonates. In this study, we investigated the genomic changes that occurred during long-term circulation of a *Kpn* ST39 clone, causing a disproportionate number of infections on the neonatal ward at a tertiary healthcare facility in Malawi in 2017.

**Methods:** We analyzed whole genome sequences of *Klebsiella pneumoniae* ST39 collected from Queen Elizabeth Central Hospital over a 20-year period, including generation of several high-quality hybrid genomes. We compared virulence markers, antibiotic resistance determinants, and mobile genetic elements, focusing on variable regions between strains from the outbreak clone in 2017 to genomes from other co-occurring ST39 lineages.

**Results:** We identified eight variable genomic regions that demonstrate the plasticity of *Kpn* within-ST, including the role of bacteriophages in shaping the genome of ST39.

**Conclusions:** The analyzed *Klebsiella pneumoniae* ST39 lineages have a highly variable genome capable of incorporating large genomic regions during prolonged hospital circulation, which may offer a selective advantage in hospital environments and provide resistance to antimicrobial agents.

**Data summary:** All sequencing data is available in BioProject PRJEB102175; detailed accession numbers are provided in Table S1. The authors confirm all supporting data, code and protocols have been provided within the article or through supplementary data files.

## Introduction

*Klebsiella pneumoniae* (*Kpn*) is a significant cause of healthcare-associated infections (HAI) globally (1, 2). HAI due to *Kpn* include lower respiratory, urinary tract, and bloodstream infections; meningitis and surgical site infections (1, 3, 4). *Kpn* infections are increasingly resistant to the World Health Organisation (WHO) recommended antibiotic regimes, and carbapenem-resistant Enterobacteriales (CRE), including *Kpn,* are designated ‘critical priority’ by the WHO (5, 6). Antibiotic resistance in *Kpn* is driven by mobile genetic elements such as transposons, bacteriophages and conjugative plasmids, as well as, less frequently, chromosomal mutations. Mobile elements can carry genes for resistance to multiple drug classes, rapidly leading to multidrug-resistant lineages (2). Mobile elements are also important for the spread of virulence determinants, which are important for host invasion, leading to clones that can cause life-threatening infections in human hosts (2). Some of the well-described virulence factors are the siderophores salmochelin, aerobactin, yersiniabactin and enterobactin, which are important for *Kpn* to cause invasive disease (7).

In low and middle-income countries, *Kpn* has emerged as a major cause of neonatal infection, and a frequent cause of outbreaks. Large multi-national studies in Africa and East Asia show that *Kpn* is the number one cause of neonatal sepsis and also the leading cause of childhood deaths (8, 9). In Malawi, more than half of all *Kpn* invasive disease in a 20-year period at Queen Elizabeth Central Hospital (QECH) in Blantyre, Malawi occurred in neonates. During this time, *Kpn* lineages have undergone continuous replacement over the years with ST15, ST14, and ST39 comprising the majority of *Kpn* invasive disease at QECH (6). These strains expand to cause outbreaks in neonatal and paediatric wards (10). However, we do not fully understand whether genomic changes take place and if these contribute to the success of specific STs in a ward environment where multiple STs are circulating (10, 11).

In this study we investigated the genomic features underlying the success of the *Kpn* ST39 lineage which caused the majority of all invasive disease in neonates at QECH in 2017. We performed a high-resolution genomic analysis including the generation of several high-quality reference genomes to explore the evolutionary mechanisms leading to the persistence of this lineage in the hospital environment.

## Methods

### Ethical permissions and approvals

This study analyses previously published data (6). Ethical approval for the parent study was granted by the University of Malawi College of Medicine Research Ethics Committee (COMREC) (P.11/18/2541).

### Samples analysed in this study

This project performs a detailed analysis of genomes previously described in a large-scale genomic epidemiology study, which revealed that the ST39 lineage caused 53.2% of invasive disease cases at Queen Elizabeth Central Hospital (QECH) in 2017 (12). QECH is one of the two tertiary hospitals in southern Malawi offering free of charge medical care to Blantyre and referrals from the surrounding districts. The ST39 genomes analyzed in this study were mainly isolated from Chatinkha nursery, which is the neonatal unit at QECH. All wards at QECH have access to a blood and cerebrospinal fluid (CSF) culture service offered by the Malawi Liverpool Wellcome Programme (MLW), who offer diagnostic services to QECH since 1998. The genomes analyzed in this study are part of the blood and CSF culture samples collected during routine clinical care of patients at QECH. We retrieved the Illumina short reads of these genomes from the European Nucleotide Archive (ENA) (supplementary table 1) (6).

### Oxford Nanopore sequencing

We selected five representative isolates 1004078, BKREGE, CAA8N9, CAAUE8, and CHI11E to use long-read sequencing on the Oxford Nanopore Technologies MinION platform for complete assemblies; the long-read sequences of CHI11E had been published already previously as part of the long-term study (6). We collected the four isolates (1004078, BKREGE, CAA8N9, CAAUE8) for long-read sequencing from the MLW bioarchive. We recovered them by plating on Nutrient agar (Thermo Scientific Oxoid, United Kingdom) and incubating under aerobic conditions at 37 for 18 hours. We transferred a single colony to 5ml of Nutrient broth (Thermo Scientific Oxoid, United Kingdom) and incubated for an additional 18 hours at 37 under aerobic conditions. We concentrated the bacterial cells from the broth by centrifugation at 3500 rpm for 20 minutes. We discarded the supernatant and extracted DNA from the pelleted cells using the MasterPure Complete DNA & RNA Purification Kit (Biosearch Technologies, United Kingdom) following the manufacturer’s instructions for extracting DNA from cultured bacterial cells. We performed quality control of the extracted DNA using the Qubit 4.0 (ThermoFisher Scientific Inc.) fluorometer and prepared the sequencing library using the SQK-RBK004 rapid library preparation kit (Oxford Nanopore Technologies plc). We loaded the library onto an R9.4.1 flow cell and sequenced the samples on the MinION Mk1c device. We acquired data from the sequencer using MinKNOW^TM^ software (Oxford Nanopore Technologies plc). We used Guppy basecalling software (version 6.0.7+c7819bc, Oxford Nanopore Technologies plc) for basecalling and demultiplexing. We ran Guppy with the default settings and the “dna_r9.4.1_450bps_sup.cfg” configuration file for basecalling using the super-accurate basecaller (sup). We passed the “SQK-RBK004” to the “--barcode_kits” and used the “--trim_barcodes” flags for the debarcoding step.

### Quality control and assembly

For the reads retrieved as above, and from SRR28748993 (CHI11E), we performed quality control on ONT reads using NanoPlot (v1.38.1) and on Illumina reads using FastQC (v0.12.1) (13, 14). Both tools were run with the default configuration. We trimmed and filtered the ONT reads using Chopper (v0.9.0) with the “-q 10 -l 500” flags to keep reads longer than 500 nucleotide bases and at least a Phred quality score of 10 (14). The quality matrices of the reads used in the analysis are available in Supplementary Figure 1. We performed *de novo* assemblies of samples with only Illumina reads using SPAdes (v3.15.5) (15, 16) implemented in Shovill (v1.1.0) (17). Shovill was run with the “—trim” parameter, which removes sequencing adapters from sequencing reads. We generated hybrid assemblies for the five strains where both short-read Illumina data and long-read ONT data were available using the Trycycler (v0.5.4) assembly pipeline (18). We generated the initial assemblies for Trycycler using Flye (v2.9.1-b1780) with the “--nano-hq” flag, Raven (v1.5.0) and MiniPolish (v0.1.2) (19, 20), (21). After running Trycycler, we polished the consensus genome using Medaka (v1.11.3) with the model r941_min_sup_g507 passed to the “-m” flag (22). We performed further polishing of the draft assemblies with Illumina reads using Polypolish (v0.5.0) (23) and then using POLCA (MaSuRCA v4.1.0) (24). We annotated all the assemblies using PROKKA (v1.14.6) with the “--usegenus --genus *Klebsiella* --addgenes --force –compliant” parameters which create Genbank compliant annotations using the *Klebsiella* specific BLAST database (25). All accessions of the hybrid assemblies and underlying reads are provided in Supplementary Table 1.

### Bacterial genome typing

To identify antimicrobial resistance genes (ARGs) and plasmid replicons we used AMRFinder Plus (v4.0.15, Database version 2024-12-18.1), with the “–plus” flag (26) and Abricate (v1.0.1) with the PlasmidFinder database (Database date 2023-11-4) to detect plasmid replicons in the assemblies (27, 28), respectively. We reconstructed plasmids from Illumina assemblies using mob_recon (v3.1.9) (29). We identified prophage regions using PHASTER and explored the identified regions using PhageScope (30–33). We used Kleborate (v3.1.3) to identify *Klebsiella pneumoniae* specific virulence factors, capsule (K-) and LPS O-antigen (O-) types (34).

### Phylogenetic analysis

We performed a whole genome core SNP analysis using Snippy (v4.6.0) with the outbreak genome BKREGE as a reference (35). We created a multiple sequence alignment of all genomes using Snippy. We identified and masked recombinant regions using Gubbins (v3.4) (36). We constructed a maximum likelihood phylogenetic tree from the recombination-free alignment with 1552 sites using IQTree (v2.2.2.7). We ran IQTree with 1000 bootstrap replicas and the General Time Reversible plus gamma (GTR+G) nucleotide substitution model from the recombinant-free alignment generated by Gubbins (36–38). We rooted the tree at midpoint using the phangorn (v2.11.1) R (v4.2.1, R Development Core Team, 2022) package and visualised it in the ggtree R package (39, 40).

To estimate the time of divergence for strains in the outbreak clade, we constructed a time-dated phylogenetic tree using the Bayesian Evolutionary Analysis Sampling Trees (BEAST,v1.10.4) software (41). We checked the alignment for a phylogenetic signal using TempEst (v1.5.3), as shown in Supplementary Figure 2 (42). BEAST was run with the GTR+G model with the empirical site heterogeneity rates, the relaxed clock model and the coalescent constant sample size option. A chain length of 800,000,000 was selected for the analysis, with logging every 5,000 states. We ran BEAST in three replicas and combined their outputs using LogCombiner (v1.10.4) to generate a maximum clade credibility tree (41).

### Statistical testing and plotting

All statistical testing was performed in the R Statistical Computing software (v4.2.1) (43). To determine whether the cases of ST39 observed in 2017 were higher than expected, we modelled the yearly counts of ST39 cases using the negative binomial model. We estimated the probability of observing the number of cases for 2017 by fitting the 2017 cases to the mean and variance of the modelled yearly counts using the’ pnbinom’ function from the’ stats’ R package (43). We used the ggpubr (v0.6.0) package to test for difference in distribution of plasmid replicons and ARGs between the different clades (44). Data wrangling and plotting were generated using the tidyverse packages (v1.3.2), ggpubr, ggblanket(v12.2.0), and patchwork(v1.2.0) (45–47).

## Results

### Outbreak in Chatinkha nursery

In a large longitudinal study analysing the collection of invasive *K. pneumoniae* isolates from QECH, we observed a marked increase in *K. pneumoniae* ST39 cases in 2017 (n = 100/188; p = 3.157208e^-11^). The number of cases later reduced to 9/208 in 2018 and 10/236 cases were observed in 2019 (Figure 1A) (48). This observed increase in cases in 2017 led us to investigate the genomes of the ST39 lineage.

**Figure 1:**
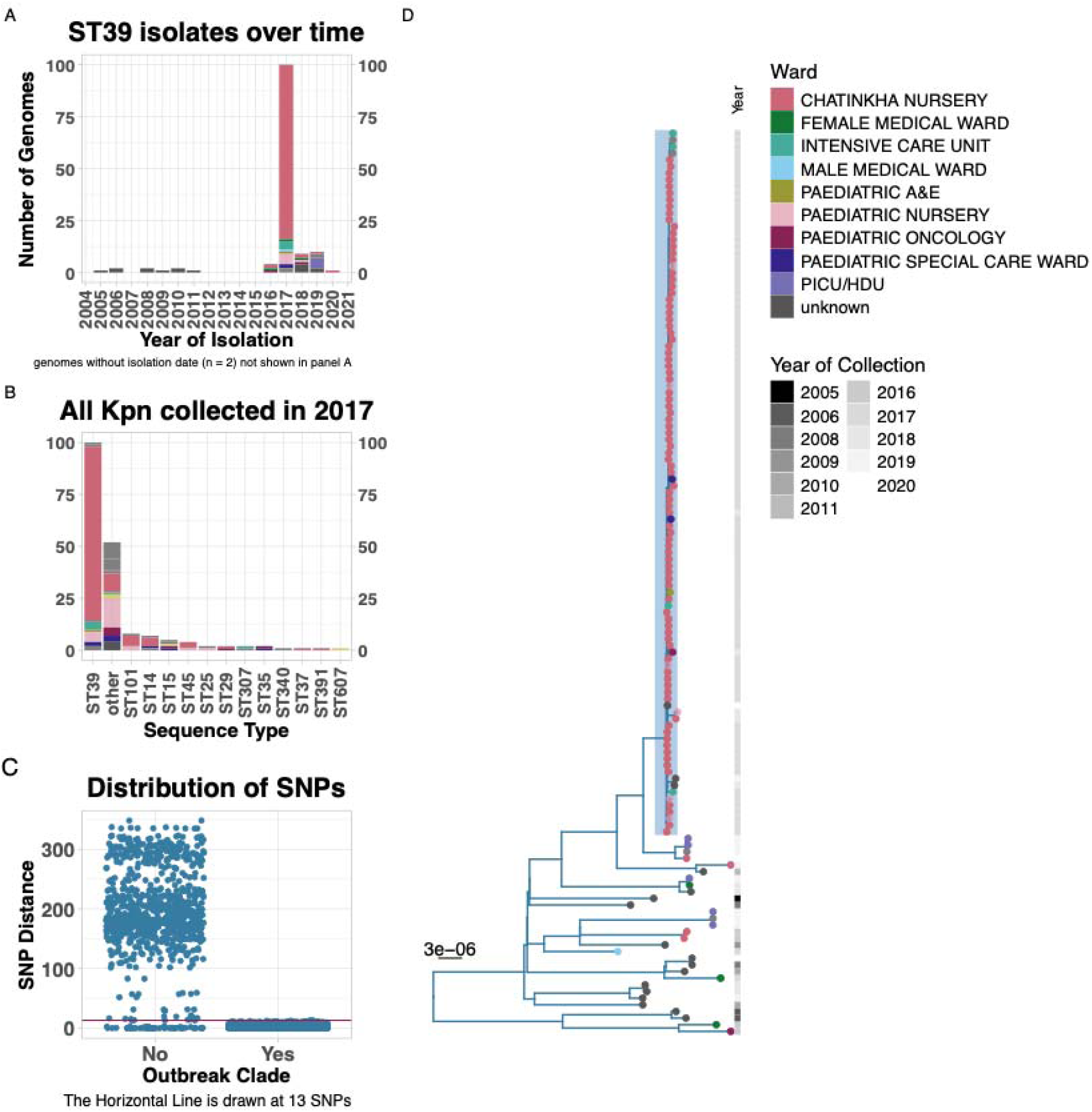
**A.** Distribution of all ST39 genomes analysed in this study across the years, the ward of sample collection is shown by colour of the bars. **B.** Distribution of all Klebsiella pneumoniae isolates collected in 2017 reported previously, highlighting the contrast in number of ST39 isolates vs other sequence types. The bars are coloured by the ward of sample collection. **C.** The distribution of pairwise SNP distances within the outbreak and non-outbreak genomes. **D.** A maximum likelihood phylogenetic tree constructed from whole genome core SNPs. The tips are coloured by the ward of sample collection, and the year of sample collection is mapped to the right of the phylogenetic tree. The isolates for which we obtained long-read sequence data are highlighted by horizontal bars from the tips of those genomes. The shaded branch represents the outbreak clade. ST, Sequence Type; SNP, Single Nucleotide Polymorphism; Paediatric E&A, Paediatric Emergency and Accident; PICU, Paediatric intensive Care Unit; HDU, High Dependence Unit.

We analysed 135 *K. pneumoniae* ST39 genomes which were collected between 2005 and 2020 (Figure 1A). The majority of the ST39 genomes were isolated in 2017 (74.1%, n = 100). The ST39 genomes from 2017 were primarily isolated from neonatal wards (87%, n = 87). This proportion contrasts with other STs, where the neonatal wards accounted for only 33% of cases of all other genomes from 2017. This was not due to differences in the numbers of samples performed on the neonatal wards in 2017, as all other STs remain in very low numbers in 2017 (overall <8 cases per ST and <6 cases among neonates) (Figure 1B).

Phylogenetic analysis revealed that 98% (n = 98) of the genomes from 2017 formed a monophyletic branch with less than 13 SNPs differentiating (median pairwise SNP distance of 2 (0 to 13)) (Figure 1C and 1D, Supplementary Figure 3). Genomes on this branch are therefore referred to as the outbreak clade genomes throughout this paper. We also observed that the genomes within the outbreak clade that were collected in 2017 all belonged to neonatal and infant wards. Only two genomes from 2017 were not included in the outbreak clade and these were collected from adult wards.

### Antimicrobial resistance genes and virulence determinants in outbreak genomes vs non-outbreak genomes

Overall, genomes in this study had a total of 47 distinct ARGs. We identified ARGs to three subclasses of beta-lactam antibiotics; beta-lactams (*bla_SHV-1_, bla_TEM-1_, bla_SHV-11_, bla_LAP-2_, bla_TEM_,* and *bla_OXA-9_*), cephalosporins (*bla_SHV_C-112A_, bla_CTX-M-15_, bla_OXA-1_,* and *bla_OXA-10_*) and carbapenems (*bla_NDM-1_).* Other classes of ARGs identified in these genomes include aminoglycoside (*aac*(*3*)*-IId, aac*(*3*)*-IIe, aph(6)-Id, aph(3’’)-Ib, aph(3’)-Ia, aadA16, aadA1, aac(6’)-Ib, aadA2,* and *aph(3’)-VI, aac(6’)-Ib-cr5*), fluoroquinolones (*qnrS1, qnrB6, qnrB1*), fosfomycin (*fosA*), macrolide (*mph(A)*), chloramphenicol (*catA1, catA2, catB3, cmlA5*), rifamycin (*arr-2, arr-3*), sulfonamide (*sul1, sul2*), tetracycline (*tet(A), tet(D)*), and trimethoprim resistance genes (*dfrA7, dfrA12, dfrA14, dfrA15, dfrA27, dfrA30*). The outbreak clade had a uniform ARG composition (Figure 2). The number of ARGs observed in the outbreak genomes was lower (p<0.0054, Wilcoxon rank sum test) than the numbers observed in the non-outbreak genomes (Figure 3A, G). The carbapenemase *bla_NDM-1_*was carried by a single genome from the non-outbreak clade (ERR12058819). This sample had six plasmids reconstructed by mob_recon, two mobilisable and four non-mobilisable. The *bla_NDM-1_* gene was located on a 98 kb IncFII(pKPX1) conjugative plasmid. The plasmid was most similar to the published plasmid sequence CP023910.1 (100% nucleotide identity, 100% query coverage) (49). This plasmid encoded for 47.3% (9/19) of the ARGs identified in this genome (*bla_NDM-1_, aac(6’)-Ib, aadA1, aph(3’)-VI, bla_CTX-M-15_, blaOXA-9, bla_TEM_, ble, qnrS1*).

**Figure 2:**
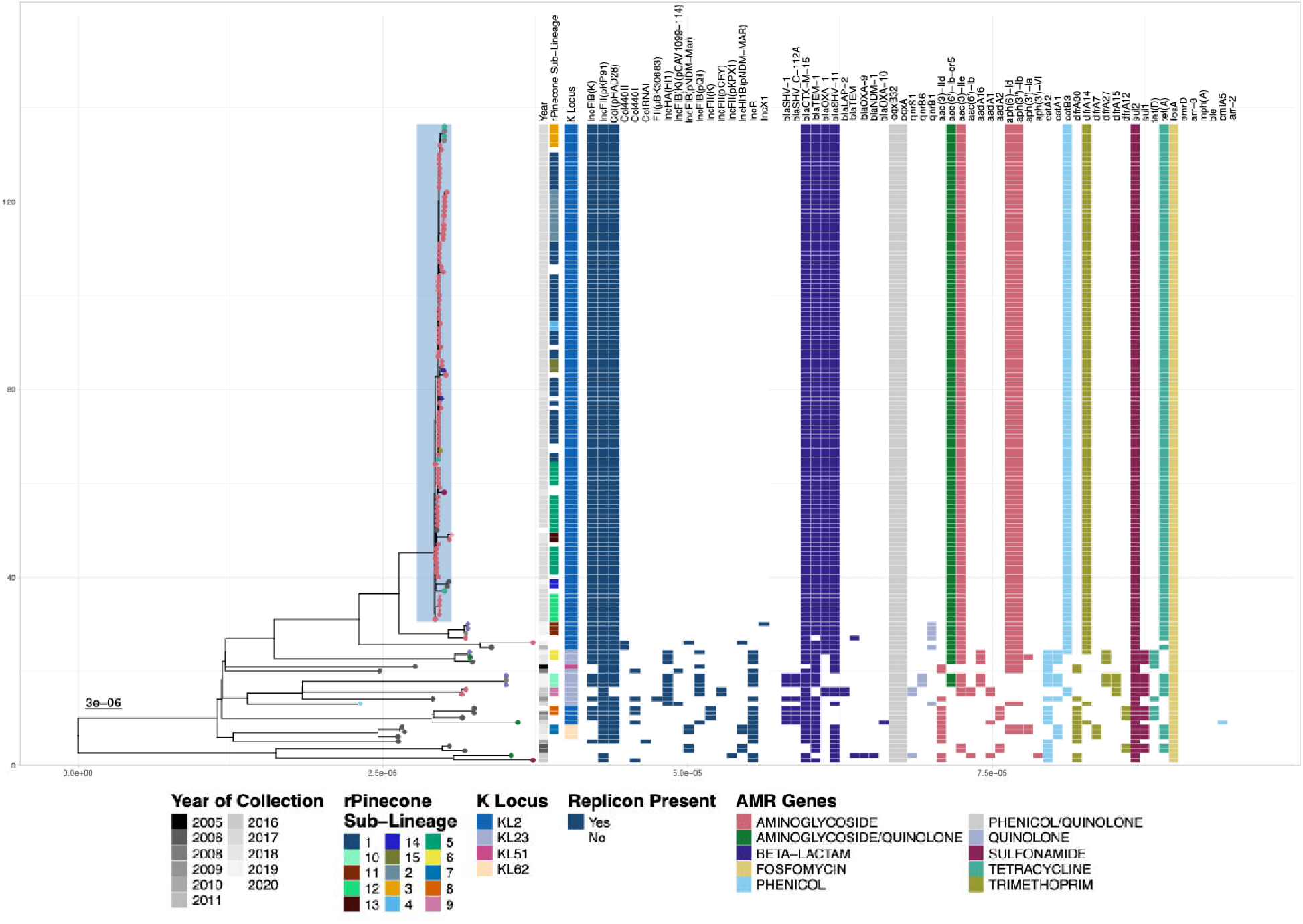
A Maximum Likelihood phylogenetic tree of all genomes in the study showing the year of isolation, sublineage by rPinecone Sub-Lineage, K Locus type, presence of plasmid replicons and antimicrobial resistance genes as a heatmap mapped to the right of the phylogenetic tree.

**Figure 3:**
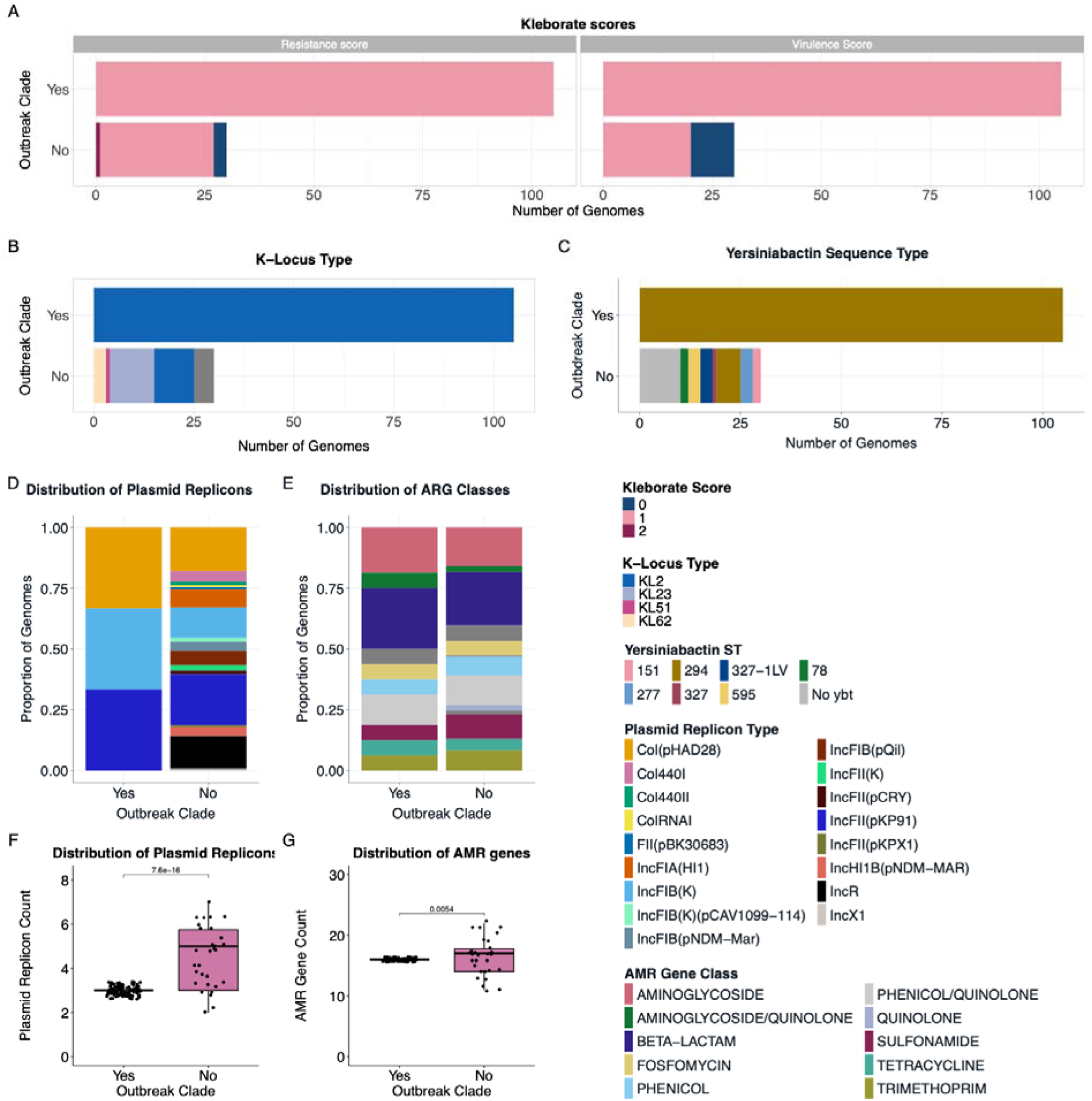
Comparison of the distribution of the number of AMR; Antimicrobial Resistance. **A.** Distribution of virulence and resistance scores calculated by Kleborate, **B.** Distribution of K-Locus types, **C.** Distribution of Yersiniabactin sequence types, **D.** Distribution of plasmid replicons as proportion of genomes and **E .** distribution of AMR gene class as a proportion of genomes. **F.** plasmid replicons per genome and **G.** ARGs identified per genome between the outbreak clade genomes and non-outbreak clade genomes. AMR, Antimicrobial Resistance; ARG, Antimicrobial Resistance Gene, ST, Sequence Type.

All genomes in this study encoded for the O1ab O-type. We noted variability in the K-types, with all isolates in the outbreak clade being K2 (KL2 locus) and other isolates having one of four other K-types (Figure 3B).

We observed a total of 17 plasmid replicon types in the collection. The genomes in the outbreak clade had a uniform composition of three replicon types including IncFIB(K), IncFII(pKP91) and Col(pHAD28). The non-outbreak genomes had varying plasmid replicon types and numbers, with a median of five (two to seven) plasmid replicon types per genome (Figure 2 and 3D). Similarly to ARGs, the number of plasmid replicons is also lower in the outbreak clade genomes (p<7.6e-16, Wilcoxon rank sum test) (Figure 3F).

### Temporal analysis using Beast

We observed that the outbreak clade shared the most recent common ancestor (MRCA) with another clade that continued to circulate in the hospital after 2017 (Figure 4). The MRCA for these 2 clades was in November 2013 (95% highest posterior density interval of March 2012 to August 2015). This divergence occurred four years before the outbreak clade genomes were isolated at QECH. The strains in the outbreak clade were isolated between March 2017 and October 2018. Multiple genomes were isolated in each month between March 2017 up to November 2017, followed by sporadic cases in 2018 (1 in January, 2 in March and another 1 in October) (Figure 4, Figures 2A and 2B). The clade that shared the MRCA with the outbreak clade continued to transmit in the neonatal wards after the outbreak. Genomes in this adjacent clade are available in our data up to April 2020. All the cases in this clade clustered together phylogenetically and form an rPinecone sublineage 8 (Figure 4).

**Figure 4:**
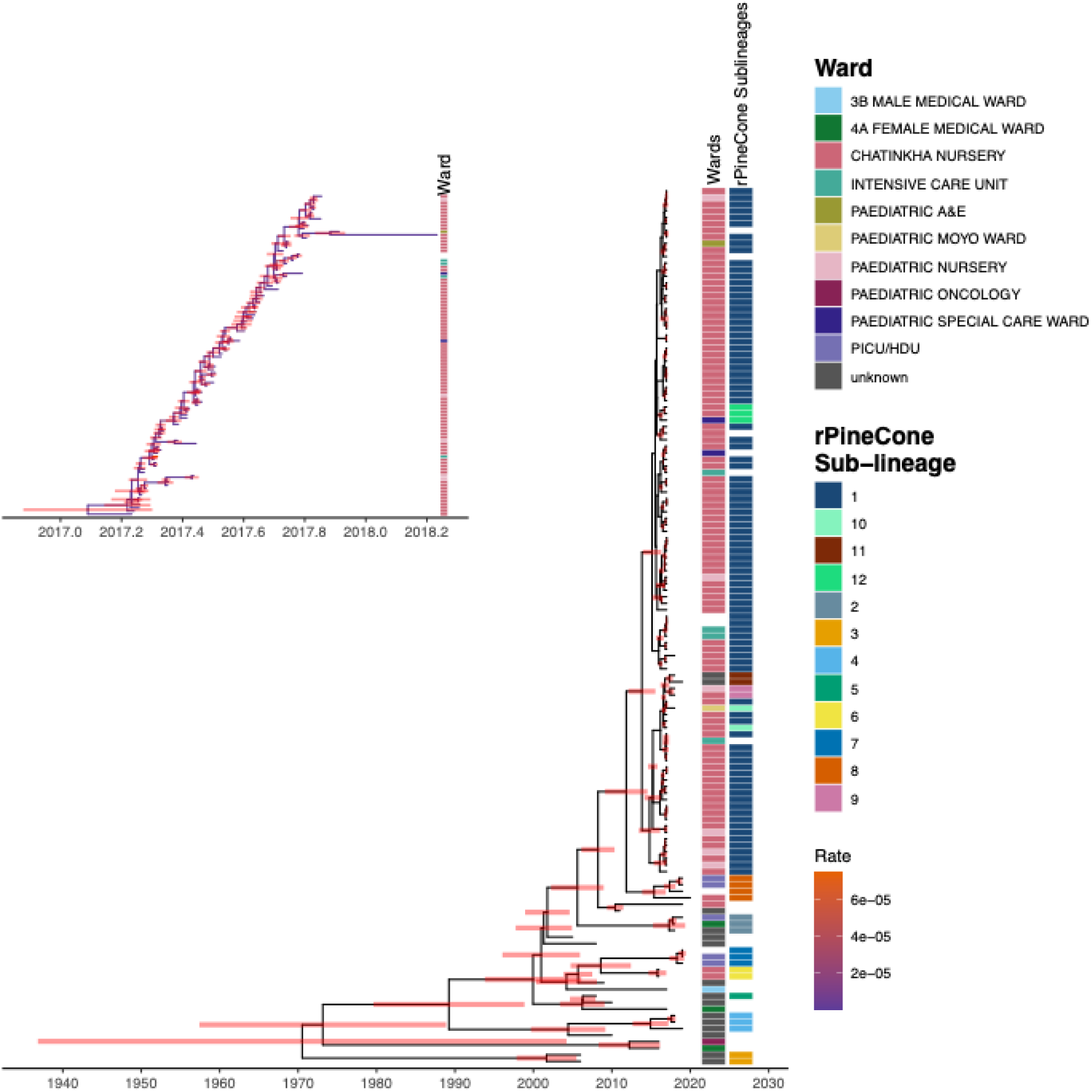
A time-dated phylogeny showing the time of divergence of the outbreak clade. The highest posterior densities are shown as bars on the nodes. The ward of isolation and rPinecone sublineages are mapped to the right of the tree. The inlet shows genomes from the outbreak clade that had complete isolation date information; 6 genomes from the outbreak clade were missing date data. The inlet has the ward of sample collection mapped to the right. The branches are coloured by branch rates. Paediatric E&A, Paediatric Emergency and Accident; PICU, Paediatric Intensive Care Unit; HDU, High Dependence Unit.

### The role of genomic plasticity in the outbreak

We identified a total of 13 variable regions when the hybrid assemblies are mapped to the reference strain BKREGE (Figure 5). Four of these regions contained prophage sequences, one was the capsule biosynthesis region and the rest had regions coding for a variety of products. Six regions (prophage 1 and 3, and region 3, 6, 7 and 8) were located next to tRNA-loci (Figure 5).

**Figure 5:**
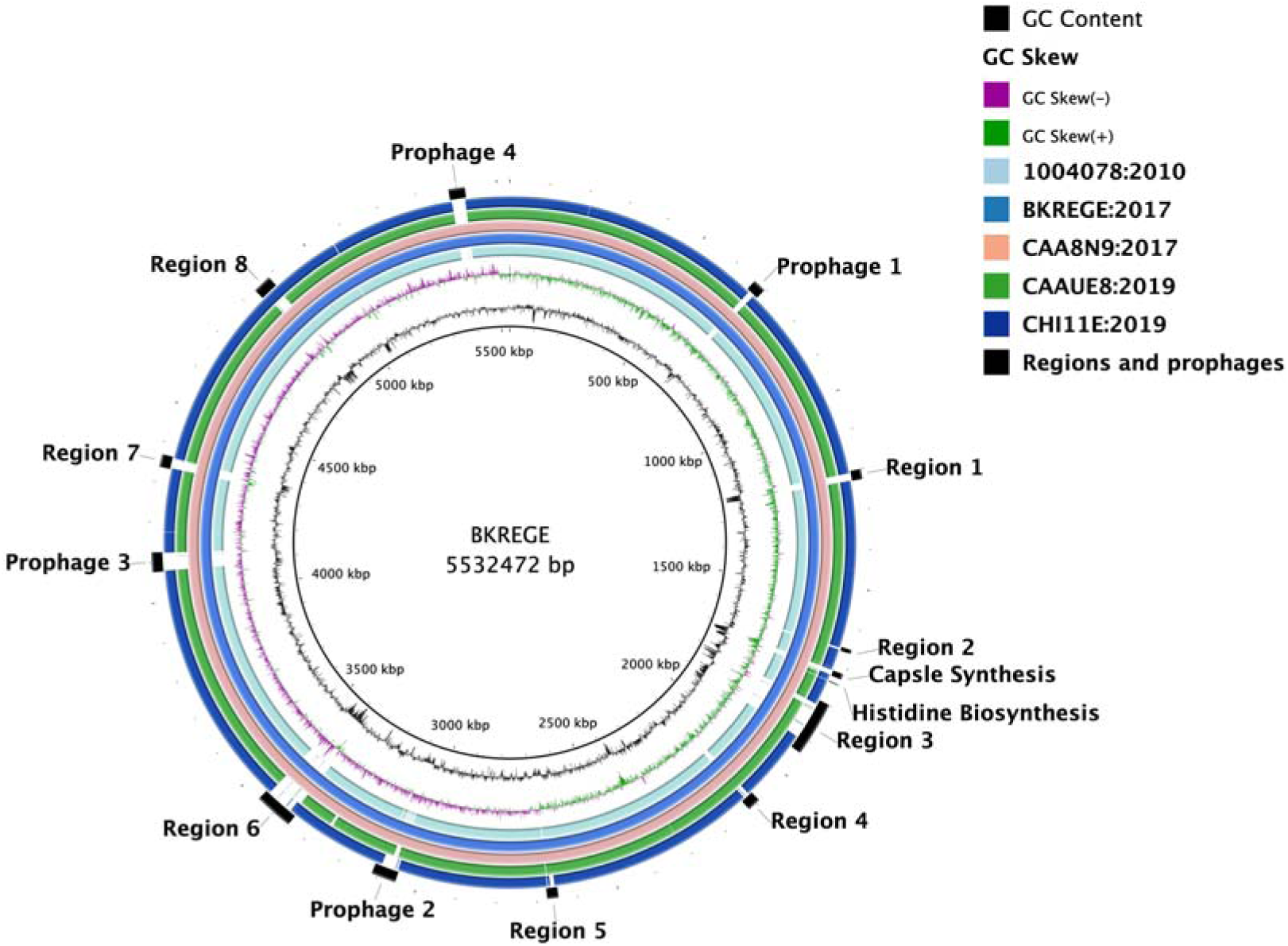
A gene map showing a comparison of chromosomes for the hybrid assemblies, highlighting prophage sequences and variable regions when the assemblies are mapped to the reference genome BKREGE. Each ring represents a chromosome in the same order as the legend.

The variable regions without prophage sequences ranged in size from 9-138 Kb (Supplementary Figure 3). While most of these regions contained coding sequences with proteins of unknown function (hypothetical proteins), three regions (variable region 3, 6 and 8) contained CDSs with known function (Supplementary Figure 3). These three regions were present in all the outbreak clade genomes but only available in a subset of the non-outbreak genomes at varying degrees (Supplementary Figure 4). Variable region 3 (Figure 5) is a 120kb region and had a type IV secretion system, CDSs involved in iron import (*irtA*), vitamin B12 import (*btuD*), and a Manganese transport operon (*mntB, mntH, mntA, mntC mntP, mntR and mntS, scaC*). Variable region 6 (Figure 5) is an 86 kb region with a toxin/anti-toxin system (*cbtA, cbtE*), antigen 43 (*flu*) that controls autoaggregation in *E.coli*, an antirestriction protein (klcA), capsular F1 antigen usher/chaperon (*caf1A and caf1M*) first isolated in *Yersinia pestis* to assist in cell surface adhesion, vitamin B12 import (*btuD*), and ferienterobactin receptor (*fepA*) (50, 51). The variable region 8 (Figure 5) is 46 Kb and mainly contains CDSs of unknown function. The region includes a toxin-antitoxin system (*ykfI, yfjZ*), an anti-restriction protein (*klcA*) and a type-1 restriction enzyme protein (*hsdS*).

We assessed each hybrid assembly for prophage sequences and identified 18 prophage regions in total. The prophage regions ranged in size from 6kb to 56 kb, with the smaller ones being from the Inoviridae family of phages (n =2, 6kb and 9kb) (Figure 5, Supplementary Figure 6. Each hybrid assembly encoded for four prophage sequences, except for isolate CAAUE8, which had only two prophages in its chromosome (Supplementary Figure 6).

Two prophage regions were conserved in multiple genomes (Figure 5). The prophage region 1 was conserved in 1004878, BKREGE and CAA8N9. The same applies to the prophage region 2, which was conserved at region 2 in 1004078 and CHI11E, and at region 1 in CAAUE8 (Figure 5, Supplementary Figure 7). The latter prophage sequence carried the *oqxAB* operon, which encodes a multidrug efflux pump (52). Four of the five assemblies carried prophage sequences known to infect the *Klebsiella* species.

Isolate CHI11E was the only one to harbor prophage regions from multiple hosts (Supplementary Figure 6). These extra prophage regions had *E. coli* and *Salmonella enterica* as their known hosts. The prophage with *S. enterica* as the known host carried a ∼50kb ARG island (Figure 6). This region is inserted between a prophage integrase (*intS)* and a *tRNA-Sec* RNA at position 96757 of the reference strain BKREGE. Similar to region 6 and 8 described above, the region has a toxin-antitoxin system, an antirestriction protein and a GTPase close to each other on one end, suggesting this section may be involved in control of transformation. The island also has the ARGs *tet(A), sul1*, *dfrA7*, and *catA1* and a mercury resistance operon (Figure 6). This insertion was seen in three genomes from the outbreak clade. Two of the three genomes were isolated in 2018, and one was isolated in 2019. These three genomes grouped into a single clade had a small genetic distance (<22 SNPs) (Supplementary Figure 8).

**Figure 6:**
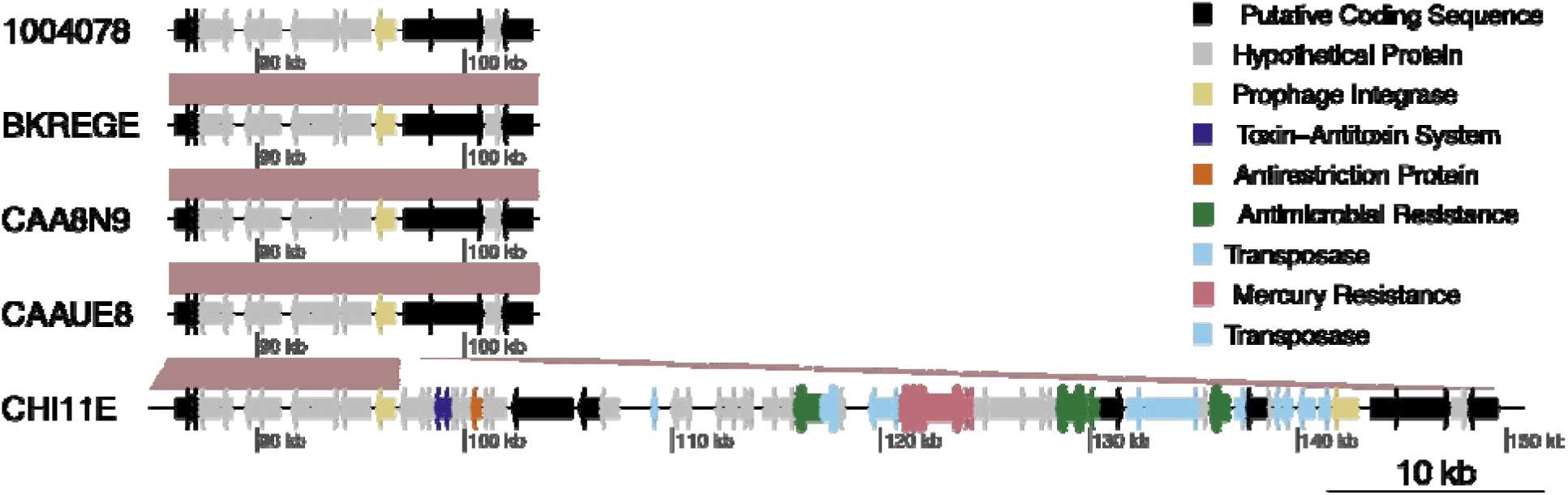
Chromosomal integration of ARGs and Mercury resistance genes. The arrows represent the coding sequences. ARG, Antimicrobial Resistance Gene.

## Discussion

Here, we performed a detailed genomic analysis of ST39 *Kpn* genomes, which caused an outbreak in a neonatal unit in Blantyre, Malawi. We utilized a combination of short and long-read sequences to perform a detailed interrogation of the genomes in the outbreak. Using a reference genome assembly from the outbreak isolates allowed us to obtain a high level of resolution, which enabled us to identify the genomic changes in this lineage. The utility of hybrid assemblies also allowed us to identify chromosomal integration of ARGs, highlighting the plasticity of AMR determinants and their impact on chromosomal structure. Our data suggest that chromosomal insertions are not randomly distributed, but rather are concentrated in certain locations along the genome of the ST39 lineage.

Our data also shows that prophage regions undergo continuous evolution, and that these regions remain important chromosomal locations for integration of future bacteriophages as highlighted by the presence of multiple prophage integrases in certain regions of the hybrid assemblies (53). These regions are also important for AMR as they can integrate cassettes carrying ARGs, which can persist and transmit vertically as observed in this analysis (54). While we are unable to show direction of evolution for these variable regions, we clearly show that they are important genomic locations for niche adaptation and AMR by the acquisition of heavy metal metabolism loci and ARGs. The presence of the insertions in genomes collected in 2018 and 2019 might indicate an incorporation and establishment of the AMR cassette in ST39. However, this phenomenon needs further investigation with more recent data, as it might have implications for bacterial fitness and survival in the hospital or human carriage niche.

The large number of insertion sequences, prophages and secretion systems identified in the variable regions suggests these regions were acquired through mobile genetic element integration into the genome of ST39. The presence of antigen 43 and fimbriae-usher encoding genes on variable region 6 of the outbreak clade suggests these genomes may have better adhesion to host cells and biofilm formation. However this needs further investigation. The genomes in the outbreak clade may have undergone similar adaptations as outlined previously (55). Such bacteria adaptation could allow the clone to have a better adaptation over other circulating strains of *Kpn*, leading to more infections.

The *bla_NDM-1_* carbapenemase identified in an isolate from this collection highlights the risk of stable introduction of carbapenem-resistant *Kpn* into Malawi, against which only very limited treatment options are available (56). The presence of distinct plasmids carrying carbapenemases in *Enterobacteriaceae* from Malawi indicates independent introductions of carbapenemases into the country (57). The plasmids carrying the carbapenemase *bla_NDM-1_*have been observed to lose this gene in absence of antibiotic pressure, leading to continued circulation of the plasmid without the carbapenemase, however, the replicon with the carbapenamase in our collection was not observed further (58). While carbapenemases are sporadically identified in isolates from Malawi, the increasing use of carbapenems at QECH might lead to selection for clones carrying these genes and lead to local circulation of the gene.

Using these ST39 genomes as an example of a lineage that expanded at QECH leading to an outbreak (6) highlights the variability within closely related *Kpn* isolates that would not be detected by core genome SNP analyses alone. A recent study from the same location observed ST39 *Kpn* in a wide range of samples, including the ward environment, indicating the ability of this lineage to persist in the environment and cause infections as ST39 and other *Kpn* STs were observed circulating in the ward environment within 28 days of either invasive disease or stool colonisation episodes (59). The isolation of our genomes in 2019 and 2020 after the ST39 outbreak matches well with the period when ST39 was observed in the environment by *Pearse et al*, further supporting the observation that invasive disease is driven by colonisation with this lineage.

Our analysis also showed that the clone implicated in the outbreak may have diverged 3-8 years from its neighbouring clades before being introduced into and isolated at QECH, suggesting possible community circulation of this lineage before it was transferred to the hospital or introduction of this clade of ST39 from other parts of the world. Regardless, our data shows the highly clonal nature of this clade, suggesting a single clone was responsible for the outbreak and may have been seeded into the neonatal unit from another point in the hospital environment. This could explain why the outbreak was interrupted somewhere in the first quarter of 2018, possibly due to the removal of the environmental reservoir. Although we were unable to ascertain the source of the outbreak in the hospital, it has been reported in our setting that *Kpn* frequently transmits between babies and mothers/guardians (60, 61). The hospital environment in the neonatal wards has also been known to be a risk factor for ESBL colonization (59). It is therefore important to disrupt the movement of *Kpn* and other pathogens through access to and implementation of appropriate IPC measures.

A limitation is the lack of clinical data to compare outcomes of infection between isolates from the outbreak clade and isolates from other ST39 lineages. The genomes are not linked to patient identifiers, and some patients may have multiple isolates, leading to an overestimation of the number of patients affected by the outbreak. Our data ends at the beginning of 2020, and we were unable to determine if the AMR insertion into the chromosome persisted in the lineage, which will be of high interest for follow-up studies to explore the fixation of *Kpn* ST39 as MDR clone.

Further work, including phenotypic studies, on strains that undergo expansion in hospital environments to understand the impact of acquired genomic material on the phenotype of the strains will be of high interest. Meanwhile, better IPC measures need to be put in place to shield vulnerable populations, including neonates, from getting exposed to *Enterobacterales* circulating in hospital environments. We also need close to real-time genomic surveillance in order to identify outbreaks early and put in place extra control measures to limit the spread of outbreak clones and protect susceptible individuals.

## Data Availability

All sequencing data is available in BioProject PRJEB102175; detailed accession numbers are provided in Table S1. The authors confirm all supporting data, code and protocols have been provided within the article or through supplementary data files.

## Acknowledgments

We acknowledge support by the MLW CORE informatics team for expert technical support. We thank the MLW clinical laboratory team and the QECH neonatal clinical team for their contributions in the generation of the original data.

## Author contributions

Conceptualization - AMZ, EH, ZAD, NAF; Data curation - AMZ, OP; Formal analysis - AMZ; Funding acquisition - EH, NAF; Investigation - AMZ, OP; Methodology - AMZ, DD, PM, EH; Project administration - EH, NAF; Resources - KK, NAF; Software - AMZ, DD, PM, EH; Supervision - EH, ZAD, PM, NAF; Validation - AMZ; Visualization - AMZ; Writing-original draft - AMZ, EH; Writing-reviewing and editing - all authors.

