## Supplementary_figures_tables for "Temporal genome dynamics of ST39 *Klebsiella pneumoniae* in a neonatal unit in Blantyre, Malawi"

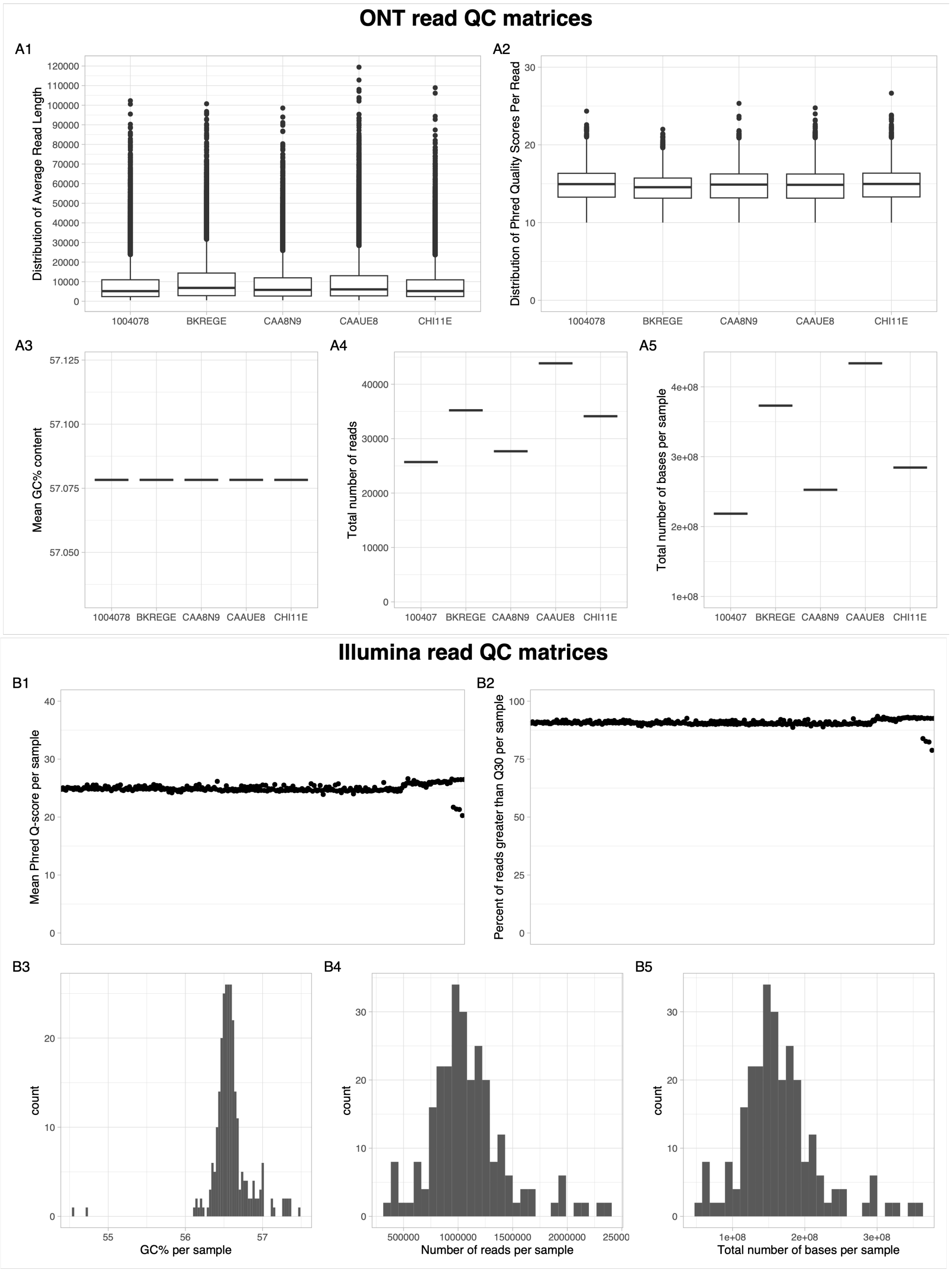


***Supplementary figure 1**: **A** Long read quality matrices. **B** Illumina read quality matrices*

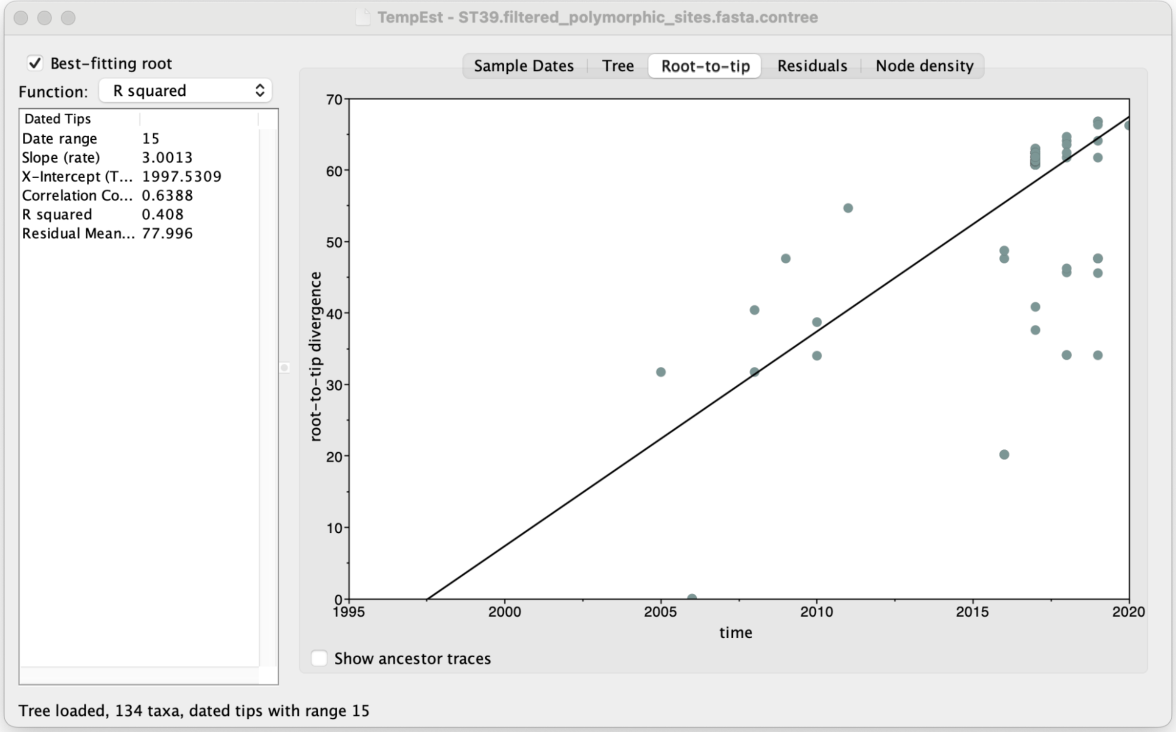


***Supplementary Figure 2:*** *TempEst output showing a root-to-tip regression plot for genomes used in the temporal analysis.*

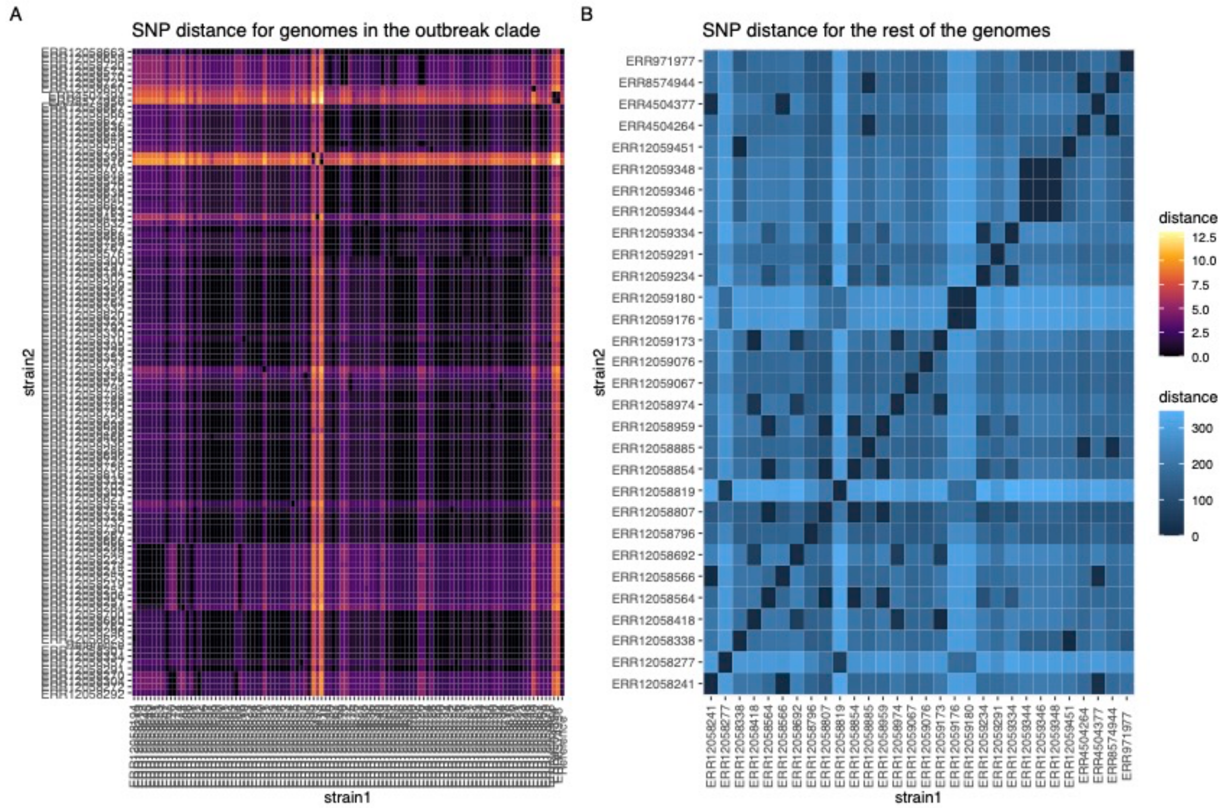


***Supplementary Figure 3***: *Pairwise SNP distances between genomes within the outbreak clade (A) and the non-outbreak genomes (B).*

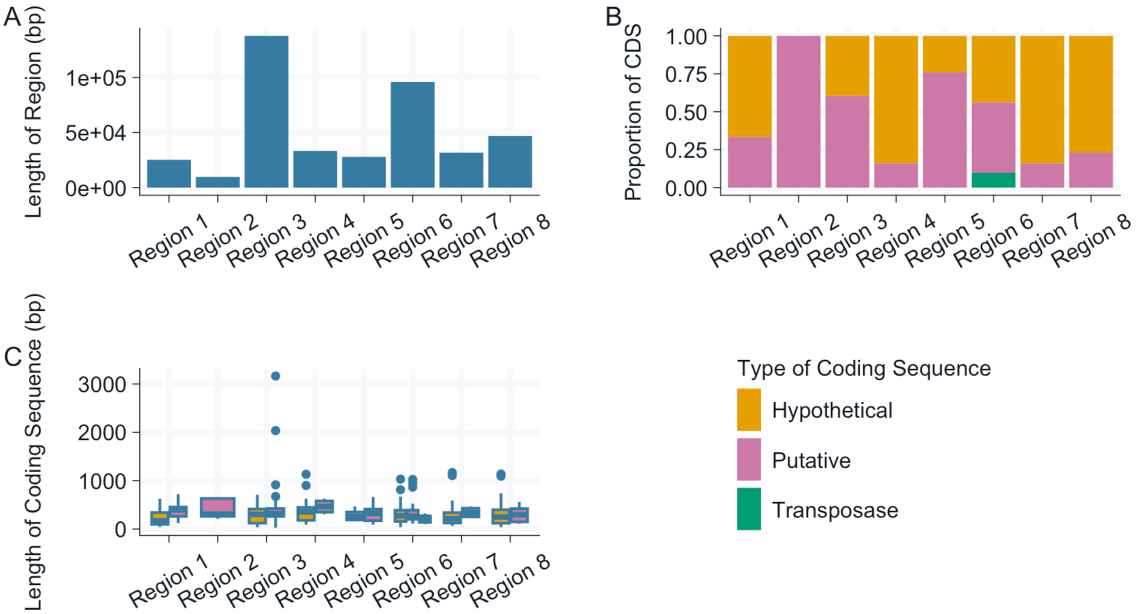


***Supplementary Figure 4:*** *Distribution of coding sequences in the variable regions assessed by length (A), proportion of elements of unknown function based on annotation (B) and length of the coding sequences (C).*

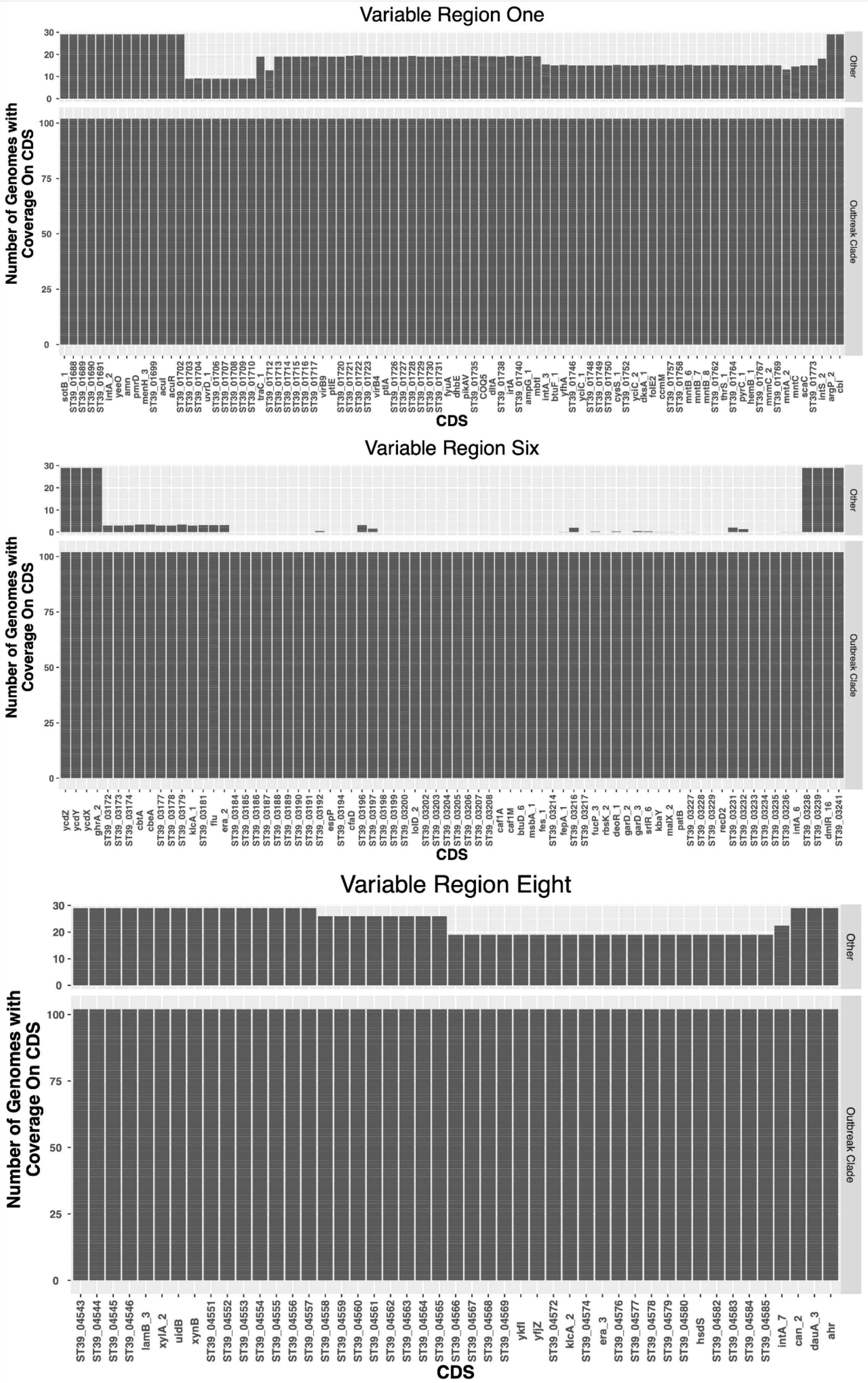

Variable Region One
Number of Genomes with Coverage On CDS
Other
Outbreak Clade
CDS
sotB_1
ST39_01688
ST39_01689
ST39_01690
ST39_01691
intA_2
yeeO
amn
pmrD
menH_3
ST39_01699
acuI
acuR
ST39_01702
ST39_01703
ST39_01704
uvrD_1
ST39_01706
ST39_01707
ST39_01708
ST39_01709
ST39_01710
traC_1
ST39_01712
ST39_01713
ST39_01714
ST39_01715
ST39_01716
ST39_01717
virB9
ptlE
ST39_01720
ST39_01721
ST39_01722
ST39_01723
virB4
ptlA
ST39_01726
ST39_01727
ST39_01728
ST39_01729
ST39_01730
ST39_01731
fyuA
dhbE
pikAV
ST39_01735
COQ5
dltA
ST39_01738
irtA
ST39_01740
ampG_1
mbtI
intA_3
btuF_1
yfhA
ST39_01746
yciC_1
ST39_01748
ST39_01749
ST39_01750
cysS_1
ST39_01752
yciC_2
dksA_1
folE2
ccmM
ST39_01757
ST39_01758
mntB_6
mntB_7
mntB_8
ST39_01762
thrS_1
ST39_01764
pyrC_1
hemB_1
ST39_01767
mnmC_2
ST39_01769
mntA_2
mntC
scaC
ST39_01773
intS_2
argP_2
cbl
Variable Region Six
Number of Genomes with Coverage On CDS
Other
Outbreak Clade
CDS
ycdZ
ycdY
ycdX
ghrA_2
ST39_03172
ST39_03173
ST39_03174
cbtA
cbeA
ST39_03177
ST39_03178
ST39_03179
klcA_1
ST39_03181
flu
era_2
ST39_03184
ST39_03185
ST39_03186
ST39_03187
ST39_03188
ST39_03189
ST39_03190
ST39_03191
ST39_03192
espP
ST39_03194
cfaD
ST39_03196
ST39_03197
ST39_03198
ST39_03199
ST39_03200
lolD_2
ST39_03202
ST39_03203
ST39_03204
ST39_03205
ST39_03206
ST39_03207
ST39_03208
caf1A
caf1M
btuD_6
msbA_1
fes_1
ST39_03214
fepA_1
ST39_03216
ST39_03217
fucP_3
rbsK_2
deoR_1
garD_2
garD_3
srlR_6
kbaY
malX_2
patB
ST39_03227
ST39_03228
ST39_03229
recD2
ST39_03231
ST39_03232
ST39_03233
ST39_03234
ST39_03235
ST39_03236
intA_6
ST39_03238
ST39_03239
dmlR_16
ST39_03241
Variable Region Eight
Number of Genomes with Coverage On CDS
Other
Outbreak Clade
CDS
ST39_04543
ST39_04544
ST39_04545
ST39_04546
lamB_3
xylA_2
uldB
xynB
ST39_04551
ST39_04552
ST39_04553
ST39_04554
ST39_04555
ST39_04556
ST39_04557
ST39_04558
ST39_04559
ST39_04560
ST39_04561
ST39_04562
ST39_04563
ST39_04564
ST39_04565
ST39_04566
ST39_04567
ST39_04568
ST39_04569
ykfI
yfjZ
ST39_04572
klcA_2
ST39_04574
era_3
ST39_04576
ST39_04577
ST39_04578
ST39_04579
ST39_04580
hsdS
ST39_04582
ST39_04583
ST39_04584
ST39_04585
intA_7
can_2
dauA_3
ahr

***Supplementary figure 5:*** *Variable presence of coding sequences in the genomes. Each bar shows the sum of read mapping to the coding sequence of the reference genome BKREGE in variable region one (top panel), six (middle panel) and eight (lower panel).*

4A

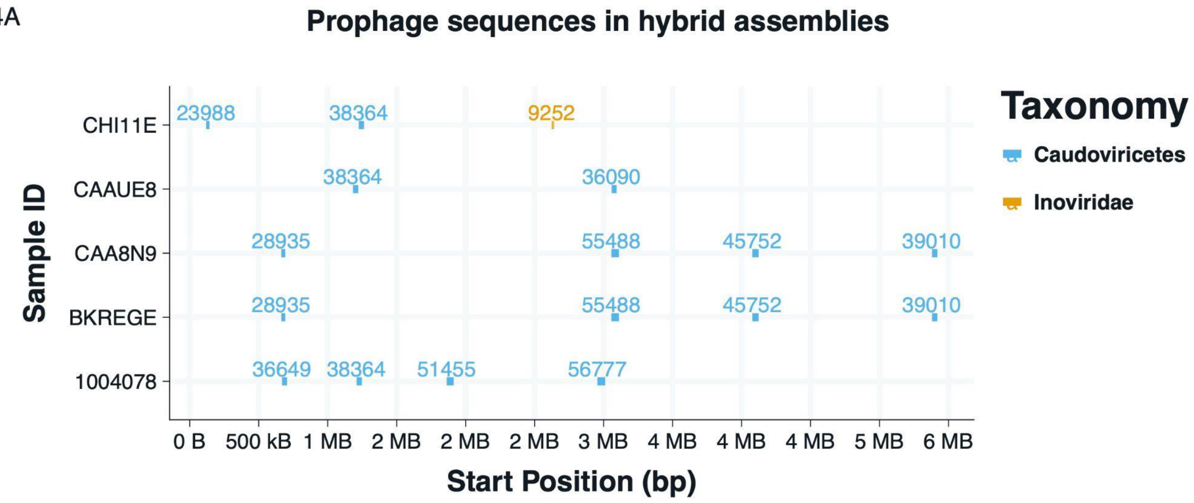


***Supplementary Figure 6:*** *The location of prophage sequences in the hybrid assemblies. The width of the bar indicates the start and end position of the prophage, the number on the bar indicates the size of the prophage sequence in base pairs, and the colour represents the virus family of the prophage sequence.*

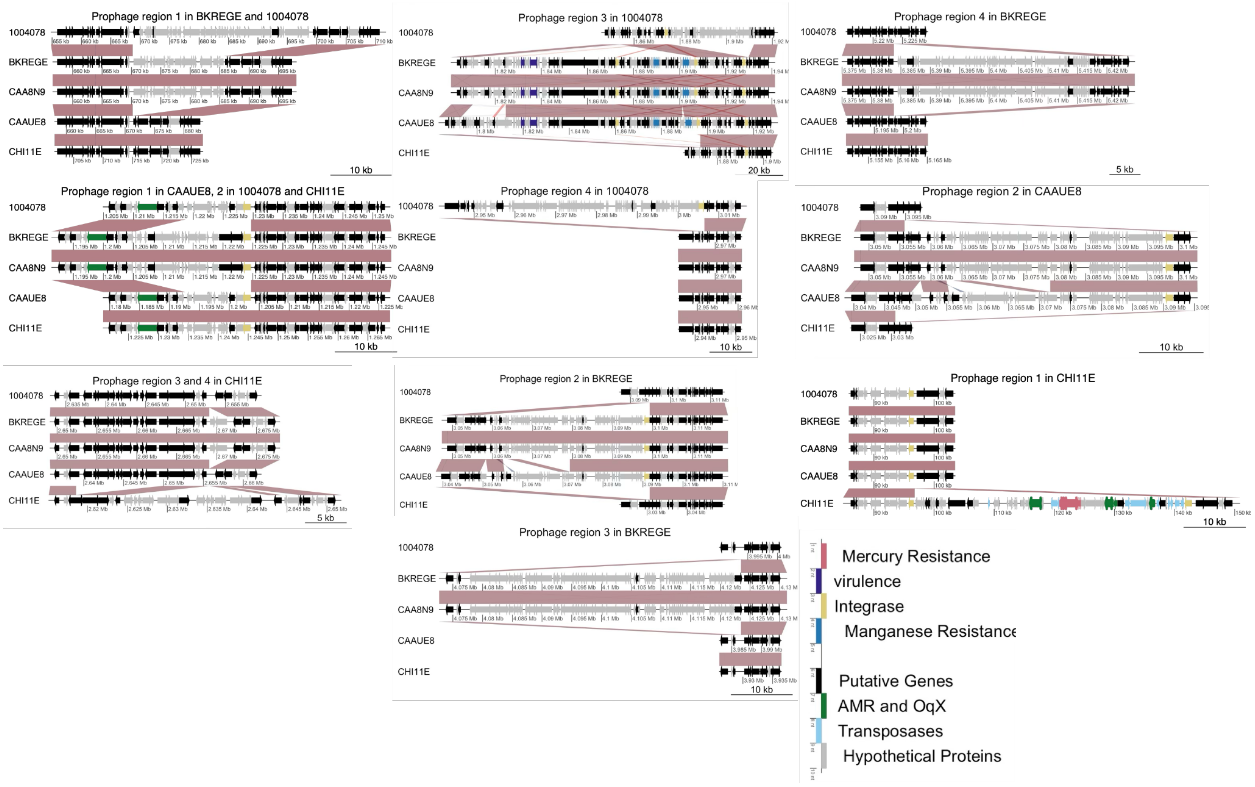


***Supplementary Figure 7:*** *Comparative analysis of prophage sequences in the hybrid assemblies.*

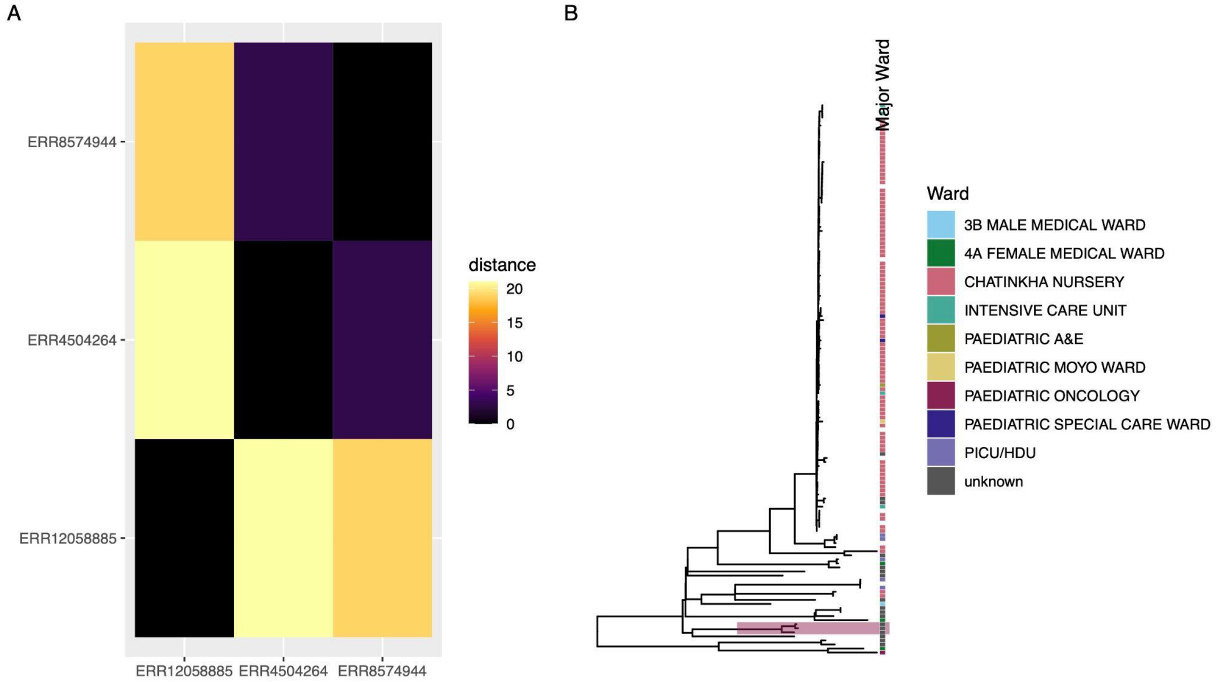


***Supplementary Figure 8:*** *Pairwise distances (A) and maximum likelihood phylogenetic tree (B) highlighting the clade with the AMR insertion.*

***Table S1.*** *Accession numbers for hybrid assemblies.*

| StrainID | Year of isolation | Clade | Illumina accession | ONT accession | Assembly accession |
|---|---|---|---|---|---|
| 1004075 | 2010 | Other | ERR12059076 | *Submitted; waiting for ENA/SRA* | GCA_977019125 |
| BKREGE | 2017 | Outbreak | ERR12058301 | *Submitted; waiting for ENA/SRA* | GCA_977019145 |
| CAA8N9 | 2017 | Outbreak | ERR12058306 | *Submitted; waiting for ENA/SRA* | GCA_977019155 |
| CAAUE8 | 2019 | Other | ERR12059348 | *Submitted; waiting for ENA/SRA* | GCA_977019135 |
| CHI11E | 2019 | Other | ERR12058885 | SRR28748993 | GCA_977019965 |
